# Cardiovascular events in individuals with small/medium LDL particle discordance

**DOI:** 10.64898/2026.06.25.26356542

**Authors:** A. Floriaan Schmidt, Nikita Hukerikar, Sam Quill, Marion van Vugt, Mathijs de Kleer, Marc Ditmarsch, Michael Szarek, John J. Kastelein, Kausik K. Ray, Michael H. Davidson

**Author notes:** **Corresponding author:** A Floriaan Schmidt. Joined senior authors (Kausik K. Ray, Michael H. Davidson).

## Abstract

**Aims:** Despite similar LDL-C levels, size and composition of LDL particles (LDL-P) varies widely. Among the metabolically perturbed, or those with altered function of lipid regulatory proteins, LDL-C levels mask elevated atherogenic small-medium LDL-P (S/M LDL-P). We assessed the contribution of such discordance in S/M LDL-P on major adverse cardiovascular event risk (MACE).

**Methods and results:** UK Biobank participants with Nightingale NMR metabolomics (487,521 participants), were classified as high or low cardiometabolic burden. S/M LDL-P discordance was defined as the difference between LDL-C predicted S/M LDL-P and observed S/M LDL-P. Genetic variants encoding cholesterol ester transfer protein (CETP), which regulates cholesterol-triglyceride exchange and the production of small LDL particles, were identified via whole genome sequencing. Adjusted Cox proportional hazard regression was used to estimate MACE associations. S/M LDL-P discordance showed an LDL-C and Apo-B independent association with MACE (47,935 cases), which differed by cardiometabolic burden group: hazard ratio (HR) per standard deviation 1.09 (95%CI 1.05; 1.13) and HR 1.24 (95%CI 1.21; 1.27) for low/high burden, respectively. Loss of function (LoF) *CETP* variants were strongly associated with lower levels of both S/M LDL-P and S/M LDL-P discordance. For example, the S/M LDL-P discordance effect of *CETP* LoF carriership for low/high metabolic burden, respectively, was -4.62 nmol/L (95%CI -8.40; -0.83) compared to -11.10 nmol/L (95%CI -15.57; -6.63).

**Conclusion:** S/M LDL-P discordance (overabundance) is strongly associated with MACE risk, especially in people with high cardiometabolic burden. S/M LDL-P discordance is modified by *CETP* genetic variation, suggesting a role for CETP-mediated lipid remodelling beyond LDL-C changes.

**Translational perspective:** Conventional lipid parameters such as LDL-C and apolipoprotein B may underestimate the atherogenic burden conferred by an overabundance of small and medium LDL particles, particularly in patients with diabetes, obesity, or established atherosclerotic disease. We introduce a novel measure of small/medium LDL particle (S/M LDL-P) discordance, quantifying the excess of S/M LDL-P beyond what is predicted by LDL-C alone. S/M LDL-P discordance is independently associated with time to incident MACE, especially in people with increased cardiometabolic burden. Genetic loss of function in cholesteryl ester transfer protein (*CETP*), which regulates cholesterol-triglyceride exchange and the production of small LDL particles, reduced S/M LDL-P discordance, in particular among those with metabolically perturbed states where discordance was otherwise high. Taken together, these findings provide support for the potential role of CETP inhibition, as a therapeutic strategy that may lower cardiovascular risk in part through reduction of S/M LDL-P discordance. This hypothesis is currently being evaluated with obicetrapib in the PREVAIL trial.

## Background

Very-low-density lipoprotein (VLDL) particles are assembled in the liver and secreted into the circulation, where they remodel with a reduction in triglycerides (TG) and increase in cholesterol esters to become intermediate-density lipoprotein (IDL) and ultimately low-density lipoprotein (LDL). This maturation process is tightly coupled to the metabolic state: insulin resistance promotes hepatic overproduction of VLDL and impairs its clearance, resulting in a shift towards smaller, denser, and more numerous LDL particles (LDL-P) with a comparatively depleted cholesterol content per particle in these three apolipoprotein-B (Apo-B) containing lipoproteins^1^. A key step in cholesterol-triglyceride exchange, which further contributes to this remodelling, occurs between apolipoprotein-A1 (Apo-A1) containing high-density lipoprotein (HDL) and Apo-B containing LDL particles (heterotypic transfer) and is mediated by cholesteryl ester transfer protein (CETP). CETP acts as a tunnel which facilitates the transfer of cholesterol esters from HDL, and Apo-B containing triglyceride-rich lipoproteins, in exchange for TG. This results in the generation of smaller, more dense, cholesterol-depleted LDL species and smaller, less abundant HDL that characterise the typical dyslipidaemia of insulin-resistant states^1^.

Although LDL cholesterol (LDL-C) has been the primary therapeutic lipid target in cardiovascular risk reduction^2–4^, it represents an aggregate measure of the cholesterol mass carried across a heterogeneous population of particles^5^. Nightingale nuclear magnetic resonance (NMR) spectroscopy has enabled the quantification of discrete lipoprotein subfractions, revealing that different particle classes carry distinct cardiovascular risk signals not fully captured by LDL-C or Apo-B alone^6^. Because VLDL-P, IDL-P, and LDL-P carry exactly one apolipoprotein B (Apo-B) molecule, Apo-B provides a measurement of the total atherogenic particle burden. Apo-B however, does not distinguish between the contributions of large triglyceride-rich remnants and the smaller, more numerous LDL-P that arise from insulin-resistant remodelling. Moreover, some studies have hypothesized that Apo-B loses some of its mass in these very small LDL particles, possibly because the molecule has to shed glycated moieties due to space constraints^7^. This implies that whilst Apo-B mass has proven to be superior to LDL-C in risk characterisation, aggregate Apo-B mass may not always reflect the true biological risk associated with different LDL-P subclasses.

Among individuals with increased cardiometabolic burden, LDL-C may appear normal whilst total LDL-P number is dominated by small and medium sized particles (S/M LDL-P)^8^. In the current study we sought to determine whether this discordance between LDL-C and S/M LDL-P is an important factor for major adverse cardiovascular event (MACE) risk. For this we defined S/M LDL-P discordance as the difference between observed S/M LDL-P concentration and the S/M LDL-P levels expected based on measured LDL-C. Given the relevance of CETP-mediated remodelling in LDL-P size and composition, as well as the ongoing pharmacologic intervention trials evaluating CETP inhibition, we additionally sought to determine the association of *CETP* genetic variants on S/M LDL-P concentration and S/M LDL-P discordance. For this purpose we had access to 487,521 UK biobank (UKB) participants with Nightingale NMR metabolites measured at the time of enrolment.

## Methods

### Study sample and data sources

The UKB is a population cohort study recruiting participants between 2006 and 2010 from across the four UK constituent countries^9^. Participants consented to sharing and linking their longitudinal electronic healthcare record (EHR) data, providing information on drug prescriptions, diagnoses, and medical procedures, sourced from hospital episode statistics, the general practitioner, and national death registries.

Physical examination, blood biochemistry and Nightingale NMR metabolites were measured at enrolment. For the current analysis we leveraged information on: age (years), Apo-B (mg/dL), blood glucose (mmol/L), body mass index (BMI, kg/m^2^), C-reactive protein (CRP, mg/L), hemoglobin A1c (HbA1c, mmol/mol), HDL-C (mg/dL), LDL-C (mg/dL), LDL-P (nmol/L), lipoprotein[a] (Lp[a], nmol/L), S/M LDL-P (the sum of small and medium LDL-P, nmol/L), sex (female/male), systolic/diastolic blood pressure (SBP/DBP, mm Hg), total cholesterol (mg/dL), TG (mg/dL), TG in S/M LDL-P (the sum of TG in small and medium LDL), TG/lipids S/M LDL-P (TG to total lipids percentage in small and medium LDL), and VLDL-C (mg/dL). We additionally considered the following derived measurements: “S/M LDL-P fraction” (defined as the ratio of S/M LDL-P by total LDL-P), non-HDL-cholesterol (non-HDL-C, defined as total cholesterol minus HDL-C), and remnant-cholesterol (remnant-Chol, defined as non-HDL-C minus LDL-C). As part of the standard Nightingale recommended quality control procedure we removed participants with fewer than 6 available NMR measurements. Based on the same guidance, values with a measurement below the lower limit of quantification were set to zero; see Supplementary Table S1 for the relevant UKB fields.

Using the available EHR data we defined atherosclerotic cardiovascular disease (ASCVD) as the composite of (fatal or non-fatal) coronary heart disease (CHD), ischemic stroke, and peripheral artery disease. MACE was defined as the composite of CHD death, non-fatal myocardial infarction, fatal and non-fatal ischemic stroke, and coronary revascularisation.

Type 2 diabetes was defined based on EHR diagnoses, prescription of insulin and oral glucose-lowering drugs, as well as using the enrolment HbA1c measurement (48 mmol/mol or higher). Pre-diabetes was defined as not having any type of diabetes (including type 1 and gestational diabetes) and an enrolment HbA1c between 39 and 48 mmol/mol^10^. Lipid lowering treatment (LLT) was based on a nurse-led interview conducted during enrolment, identifying people taking statins, ezetimibe, or fibrates (as monotherapy or combination); during the UKB enrolment phase PCSK9 inhibitors were not yet approved in the UK. Prescriptions of blood pressure-lowering medications (beta blockers, diuretics, calcium channel blockers, alpha blockers, alpha-2 receptor agonists, renin angiotensin aldosterone system inhibitors, vasodilators, and centrally acting antihypertensives), were similarly defined. Please see supplementary Table S2-S3 for the employed coding.

### Defining cardiometabolic burden

To explore potential difference by metabolic state all analyses were stratified by baseline cardiometabolic burden. A person was classified as “high cardiometabolic burden” if they met any of the following criteria at baseline: a history of ASCVD, any diabetes, pre-diabetes, obesity (BMI ≥ 30 kg/m²), total cholesterol ≥ 200 mg/dL, or TG ≥ 150 mg/dL. Those meeting none of these criteria were classified as “low cardiometabolic burden”.

### Genetic carriership of CETP variants

Ensembl Variant Effect Predictor (VEP)^11^ was used to identify *CETP* (ENSG00000087237) variants with a predicted protein truncating (PTV) consequence likely resulting in loss of function (LoF) of CETP. Specifically, we selected *CETP* variants belonging to the canonical transcript with the following VEP predicted consequence: stop gained, stop lost, start lost, frameshift, splice acceptor, and splice donor. These were supplemented with potential PTV variants from AlphaMissense^12^; see Supplementary Table S4 for the 3,316 identified *CETP* variants.

We additionally considered three *CETP* variants associated with increased CETP activity (GRCh38 chr:pos:ref:alt): 16:56983407:G:A, 16:56981179:G:C, and 16:56982180:G:A, with the first variant listed as a gain-of-function (GoF) variant in GoFcards^13^.

### Statistical analysis

Sourcing the subset of people with low cardiometabolic burden we regressed S/M LDL-P on LDL-C using a linear regression model. Subsequently, S/M LDL-P discordance was defined as the difference between observed S/M LDL-P and LDL-C predicted S/M LDL-P (i.e. the model residuals), applying the model coefficients in the entire UKB sample irrespective of their cardiometabolic burden; see Supplementary Table S5 for the regression coefficients.

A linear regression model was used to estimate the associations of genetics variants in *CETP,* and HDL-C (the canonical downstream effect of *CETP,* proxying CETP activity), S/M LDL-P, as well as S/M LDL-P discordance. To prevent potential population stratification bias genetic analyses were conducted in people of European ancestry, and additionally adjusted for the first 6 genetic principal components. Carriership of *CETP* PTVs was treated as a binary exposure, whereas the three CETP activity-increasing variants were modelled using a standard additive dosage model, counting the number of effect alleles (i.e. 0, 1, or 2).

Cox proportional hazards (PH) regression was used to estimate the associations between the lipid variables per standard deviation (SD) increase and time till MACE onset or censoring (due to end of study, non-CV death, or loss-to-follow-up), specifically considering: LDL-C, VLDL-C, non-HDL-C, remnant-Chol, Apo-B, LDL-P, S/M LDL-P, S/M LDL-P discordance. Time-to-event was determined as the time since enrolment and diagnosis, death, end of study, or lost-to-follow-up; whichever occurred first. Models were corrected for baseline sex, age, BMI, SBP, Lp[a], HbA1c, glucose, CRP, baseline LLT, and blood pressure lowering treatments.

We additionally explored mediation of the S/M LDL-P discordance association with time till MACE. Here the unmediated total effect of S/M LDL-P discordance on MACE was compared to the direct effect of S/M LDL-P discordance which remained after adding total TG, TG in S/M LDL-P, or TG/lipids S/M LDL-P, in the covariate adjusted Cox regression model.

### Sensitivity analyses

Throughout, models were stratified by high or low cardiometabolic baseline status, with interaction tests used to evaluate differences between subgroups^14,15^. The proportional hazards assumption was evaluated by correlating Schoenfeld residuals against follow-up time, finding no meaningful violations.

We additionally determined the Spearman correlation between NMR derived LDL-C, Apo-B, and TG and their directly assayed biochemistry counterparts. To evaluate the robustness of S/M LDL-P discordance, a biochemistry-based discordance variable was additionally derived by regressing NMR-derived S/M LDL-P on biochemistry-measured LDL-C. Associations between this biochemistry-derived S/M LDL-P discordance and *CETP* variant carriership, as well as time to MACE onset, were subsequently confirmed.

The limited missing data were imputed using sklearn’s K-nearest-neighbours algorithm, with 50 neighbours. Results are presented as mean differences or hazard ratios (HRs) with 95% confidence intervals (CI), and where appropriate p-values for two-sided hypothesis tests^16^. Analyses were conducted using Python 3.11, and the packages statsmodels^17^, sklearn^18^, lifelines^19^, and plot-misc.

### Ethical approval

The UK Biobank has received ethical approval from the Northwest Multi-centre Research Ethics Committee (MREC) as a Research Tissue Bank (RTB) approval. All participants provided informed consent. This study was conducted under the UK Biobank Resource Application Number 12113.

## Results

NMR measurements were available from 487,521 UK Biobank participants, of which 309,355 belonged to the high cardiometabolic burden group and 178,166 to the low burden group. People in the high cardiometabolic burden group were more likely to be male (143,665, 46%, compared to 79,334, 45%) and were older (57.59 years, SD: 7.69, compared to 54.69 years, SD: 8.44); Supplementary Table S6. The high cardiometabolic burden group had higher concentrations of total TG and Apo-B containing particles (including LDL-C and S/M LDL-P), and showed increased concentrations of blood glucose, HbA1c, and CRP; Figure 1. Across a median follow-up of 13.5 years (quartile 1, Q1 12.6; Q3 14.3) 47,935 (9%) individuals developed incident MACE, of which 35,129 (11% of 309,355) occurred in the high cardiometabolic group.

**Figure 1.**
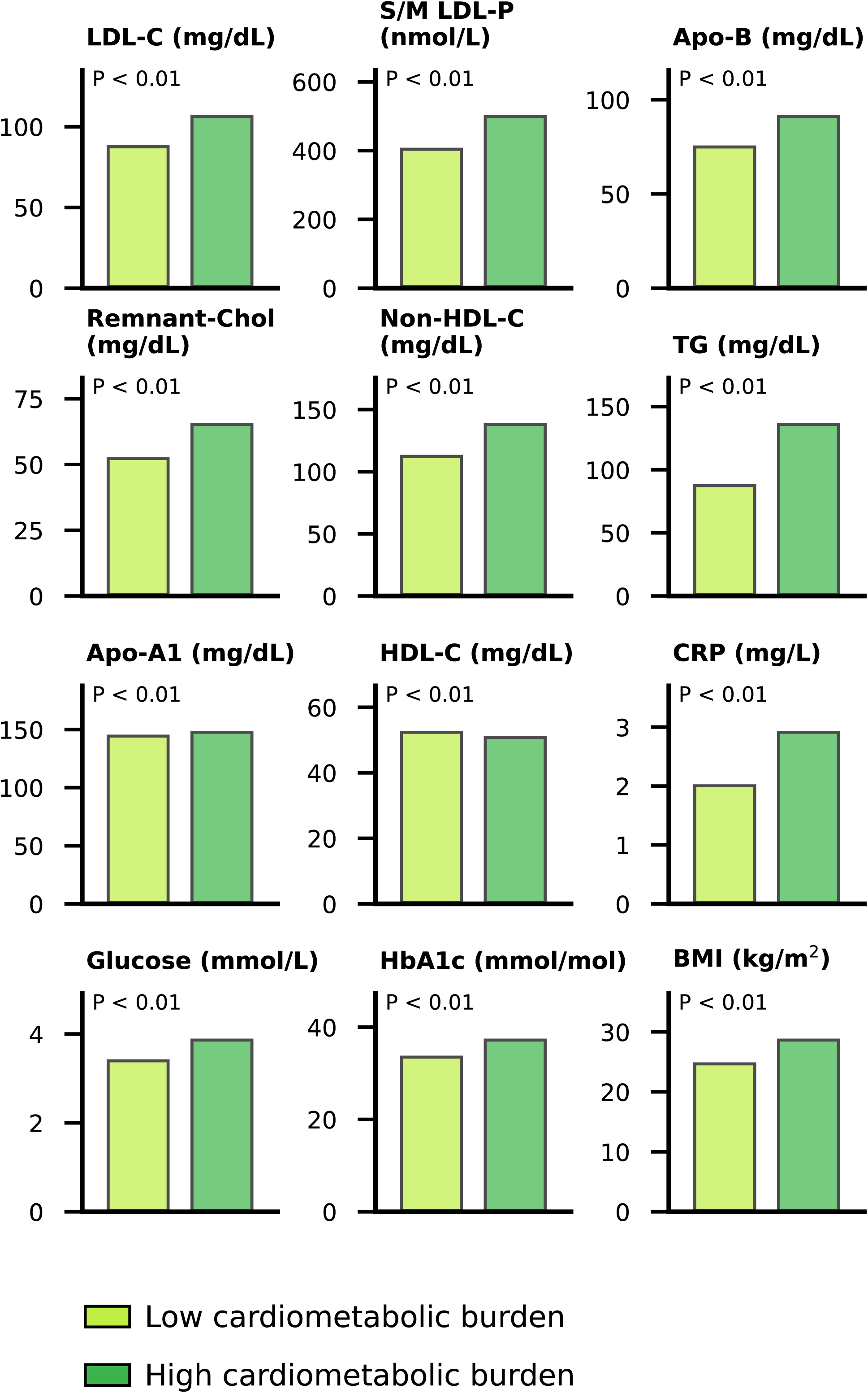
Baseline difference between UK biobank participants stratified by baseline cardiometabolic burden status. N.b. Results are based on the 487,521 UK biobank participants with NMR metabolites measurements which passed standard quality control steps. Differences between cardiometabolic groups were evaluated using a Mann-Whitney U test. Low or high cardiometabolic baseline status was defined based on the presence or absence of a history of ASCVD, any diabetes, pre-diabetes, obesity (BMI ≥ 30 kg/m²), a total cholesterol ≥ 200 mg/dL, or a total triglycerides ≥ 150 mg/dL. Abbreviations: Apo, apolipoprotein; BMI, body mass index; CRP, C-reactive protein; HbA1c, hemoglobin A1c; HDL, high-density lipoprotein; LDL, low-density lipoprotein; LDL-P, low-density lipoprotein particle; S/M, small/medium; TG, triglycerides. Please see Supplementary Table S6 for the underlying data.

### Comparing S/M LDL-P to S/M LDL-P discordance

S/M LDL-P concentration was strongly correlated with other Apo-B containing particles, LDL-C, and total Apo-B concentration; Figure 2 and Supplementary Table S7.

**Figure 2.**
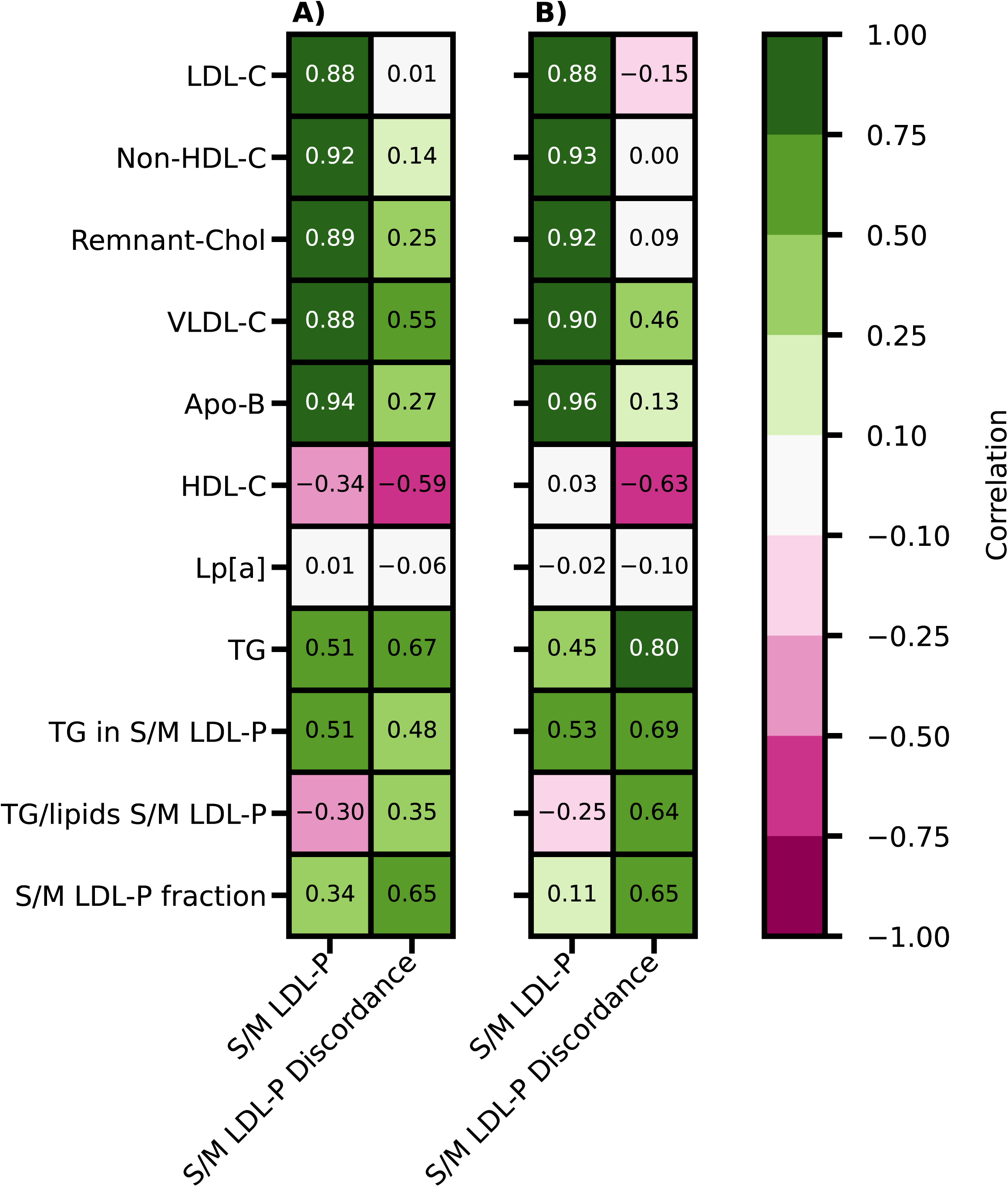
The pairwise Spearman correlation of S/M LDL-P concentration and S/M LDL-P with conventional lipid measures stratified by people with low (A) and high (B) baseline cardiometabolic burden. N.b. results are based on the 487,521 UK biobank participants with NMR metabolites measurements which passed standard quality control steps. Low or high cardiometabolic baseline status was defined based on the presence or absence of a history of ASCVD, any diabetes, pre-diabetes, obesity (BMI ≥ 30 kg/m²), a total cholesterol ≥ 200 mg/dL, or a total triglycerides ≥ 150 mg/dL. Abbreviations: Apo, apolipoprotein; HDL, high-density lipoprotein; LDL, low-density lipoprotein; LDL-P, low-density lipoprotein particle; Lp[a], lipoprotein a; S/M, small/medium; TG, triglycerides; VLDL, very low-density lipoprotein. Please see Supplementary Table S7 for the underlying data.

By contrast S/M LDL-P discordance (reflecting overabundance of S/M LDL-P) was minimally correlated with LDL-C (0.01 in the high cardiometabolic group and -0.15 in the low cardiometabolic burden group), with similar attenuated correlations for Apo-B, non-HDL-C, Remnant-Chol, and to a lesser extent VLDL-C; Figure 2. Compared to measured S/M LDL-P, individuals with increased S/M LDL-P discordance, not only had higher concentrations of total TG but also hadqualitative changes with higher TG in S/M LDL-P, increased TG/lipids S/M LDL-P, as well as an increased S/M LDL-P fraction (correlations: 0.65 in both cardiometabolic subgroups) – meaning that these discordant individuals had a higher number of S/M LDL-P compared to the total LDL-P number and these particles were enriched for TG content. Both S/M LDL-P and S/M LDL-P discordance were minimally correlated with Lp[a]; Figure 2.

### The association of genetic variants in CETP with HDL-C, S/M LDL-P, and S/M LDL-P discordance

As a positive control step we confirmed that carriership of a *CETP* LoF variant was associated with a strong increase in HDL-C concentration (Figure 3): mean difference 7.05 nmol/L (95%CI 5.79; 8.31, p-value 6.45×10^-2^) for the low cardiometabolic burden group, compared to a mean difference of 9.74 nmol/L (95%CI 8.64; 10.84, p-value 1.44×10) for high cardiometabolic burden group, respectively. Similarly, *CETP* variants associated with increased CETP activity showed the anticipated decrease in HDL-C concentration; Figure 3.

**Figure 3.**
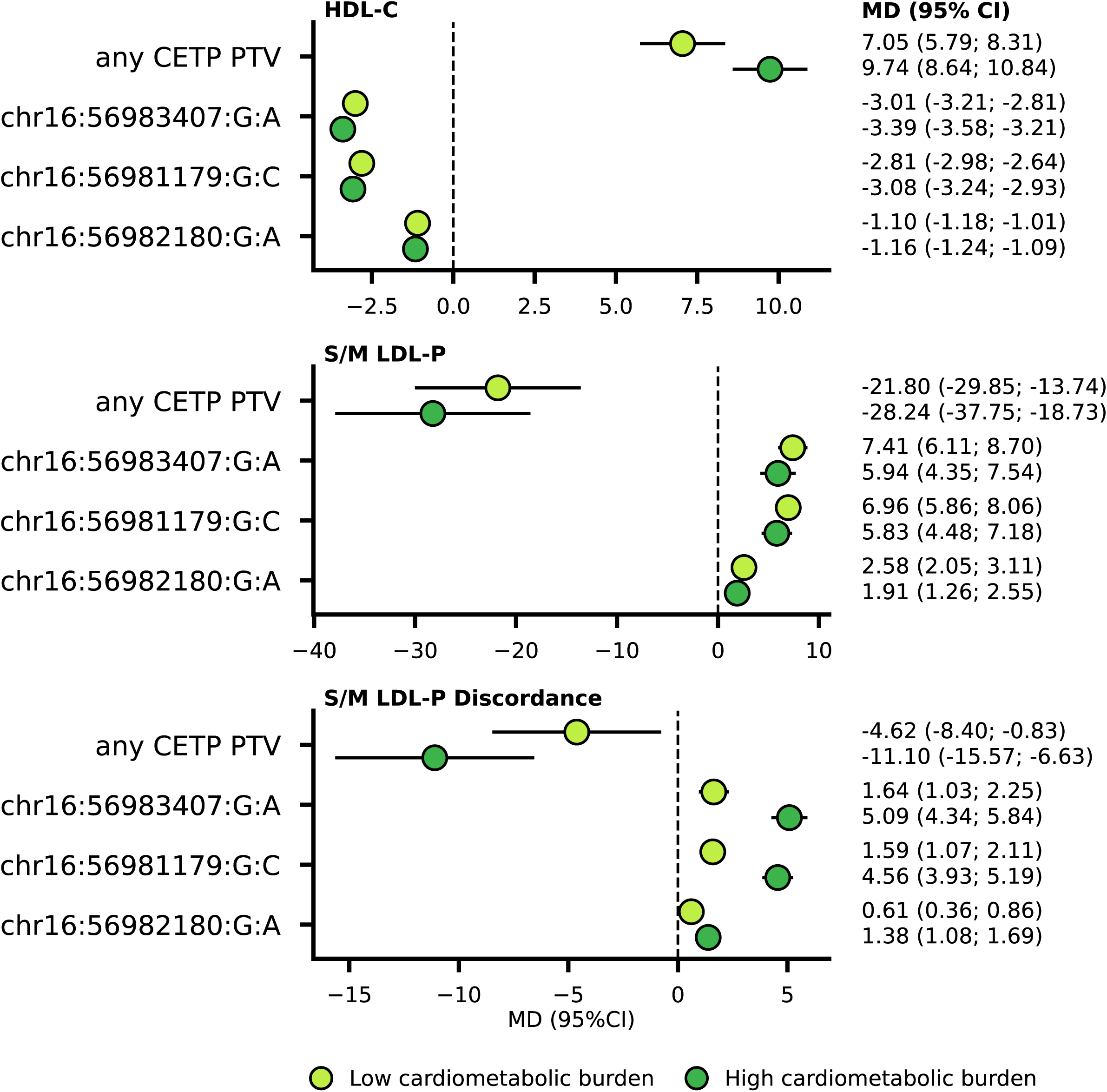
The association of genetic variants in *CETP* with HDL-C, S/M LDL-P, and S/M LDL-P discordance, stratified by baseline cardiometabolic burden status. N.b. Results are based on a linear regression model using the 387,945 UK biobank participants with NMR metabolites measurements which passed standard quality control steps. Models were adjusted for the first six genetic principal components. Out of the 141,545 European ancestry participants with low baseline cardiometabolic burden 276 carrier a *CETP* PTV, for the 246,400 European ancestry participants with high baseline cardiometabolic burden this was 516. The minor allele frequencies were 0.04/0.04 for variant 16:56983407:G:A, 0.06/0.06 for variant 16:56981179:G:C, and 0.31/0.32 for variant 16:56982180:G:A, stratified by low/high baseline cardiometabolic status. Low or high cardiometabolic baseline status was defined based on the presence or absence of a history of ASCVD, any diabetes, pre-diabetes, obesity (BMI ≥ 30 kg/m²), a total cholesterol ≥ 200 mg/dL, or a total triglycerides ≥ 150 mg/dL. Abbreviations: CI, confidence interval; CETP, cholesteryl ester transfer protein; HDL, high-density lipoprotein; LDL-P, low-density lipoprotein particle; MD, mean difference; PTV, protein truncating variants; S/M, small/medium. Please see Supplementary Table S8 for the underlying data.

*CETP* PTV carriership was associated with decreased concentration of S/M LDL-P and S/M LDL-P discordance irrespective of cardiometabolic burden; Figure 3. By contrast the three variants associated with an increased CETP activity showed a cardiometabolic burden-dependent association with higher S/M LDL-P discordance, where associations were more pronounced in the high burden group (interaction p-values < 1.30×10^-4^); Figure 3, Supplementary Table S8.

### Association of lipids measurements and incident MACE

The positive control Apo-B containing lipid measurements were all associated with time to onset of MACE, with consistent associations across cardiometabolic burden groups; Figure 4. For example, the MACE association for total Apo-B per SD was (HR 1.09, 95%CI 1.07; 1.11) and (HR 1.11, 95%CI 1.09; 1.12) for the low and high cardiometabolic burden groups, respectively.

**Figure 4.**
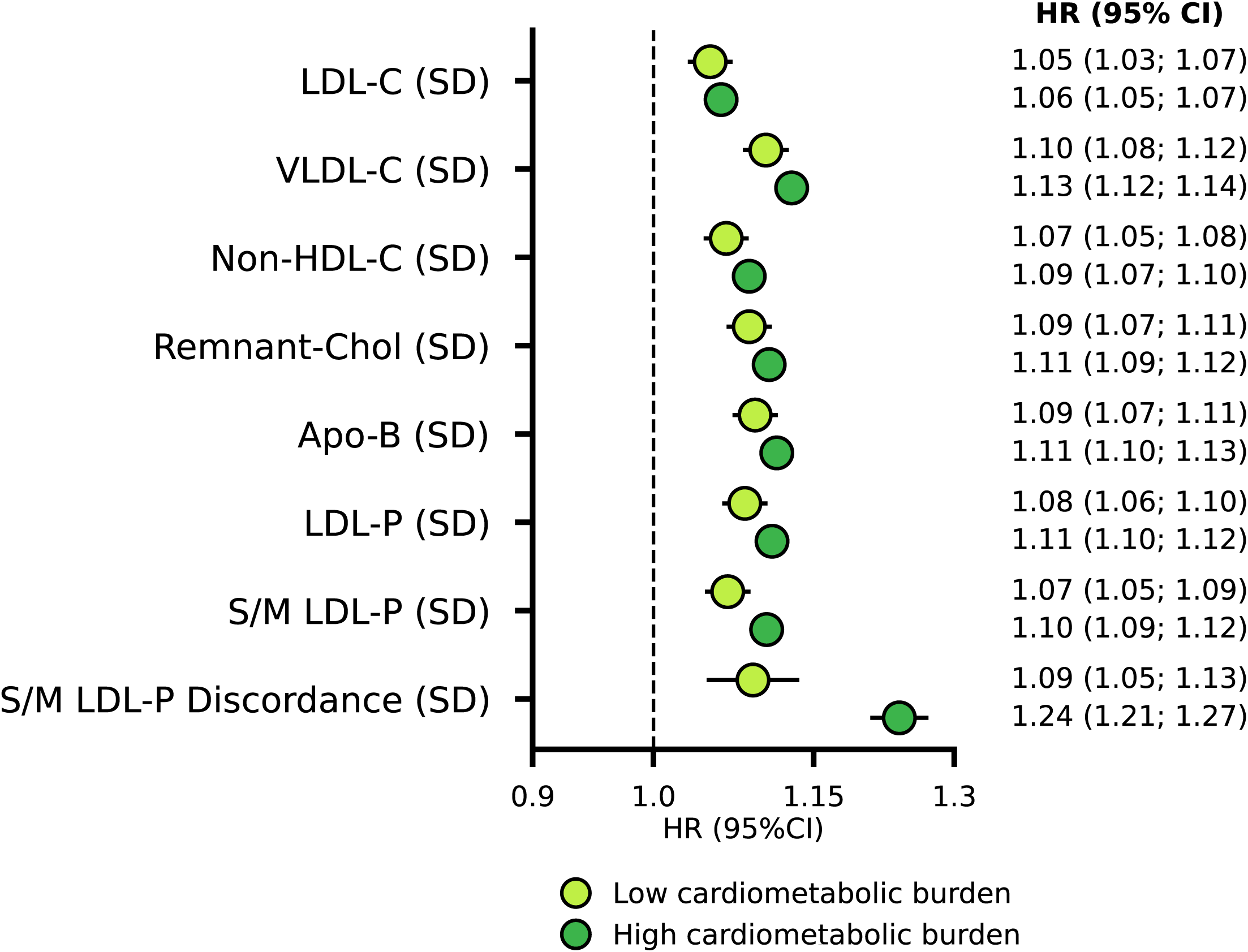
The association between lipid exposure and the time until major adverse cardiovascular events by cardiometabolic burden. N.b. Results are based on Cox regression analyses using 487,521 UK biobank participants with NMR metabolites measurements which passed standard quality control steps. Models were adjusted for sex, age, body mass index, systolic blood pressure, HbA1c, glucose, lipoprotein a, C-reactive protein, lipid lowering therapies, and blood pressure lowering treatments. Low or high cardiometabolic baseline status was defined based on the presence or absence of a history of ASCVD, any diabetes, pre-diabetes, obesity (BMI ≥ 30 kg/m²), a total cholesterol ≥ 200 mg/dL, or a total triglycerides ≥ 150 mg/dL. Estimates were standardised to an increase in exposure standard deviation, where for comparisons purposes the S/M LDL-P and S/M LDL-P discordance estimates were standardised to the same standard deviation. Abbreviations: Apo, apolipoprotein; HDL, high-density lipoprotein; HR, hazard ratios; LDL, low-density lipoprotein; LDL-P, low-density lipoprotein particle; S/M, small/medium; VLDL, very low-density lipoprotein. Please see Supplementary Table S8-S9 for the underlying data.

The HRs for total LDL-P were 1.08 (95%CI 1.06; 1.10) for the low cardiometabolic burden group and 1.11 (95%CI 1.10; 1.12) for the high cardiometabolic burden group. For S/M LDL-P the HRs were 1.07 (95%CI 1.05; 1.09) and 1.10 (95%CI 1.09; 1.12), respectively. For S/M LDL-P discordance the HR estimates were 1.09 (95%CI 1.05; 1.13) and 1.24 (95%CI 1.21; 1.27) (interaction p-value 1.19×10^-6^), stratified by low and high cardiometabolic burden groups; Figure 4 and Supplementary Tables S9-S10.

### TG mediation of the S/M LDL-P discordance association with MACE

Given the positive correlation of S/M LDL-P with total TG and S/M LDL-TG we next sought to explore potential TG-based mediation of the S/M LDL-P discordance MACE associations.

Including TG/lipids S/M LDL-P into the covariate adjusted model resulted in limited attenuation of the association between S/M LDL-P discordance with MACE. Including total TG or TG in S/M LDL-P resulted in a more meaningful attenuation; Table 1. Specifically, the TG in S/M LDL-P adjusted association of S/M LDL-P discordance on MACE was HR 1.02 (95%CI 0.98; 1.07) and 1.08 (95%CI 1.04; 1.12), for the low/high burden groups, respectively.

**Table 1.**
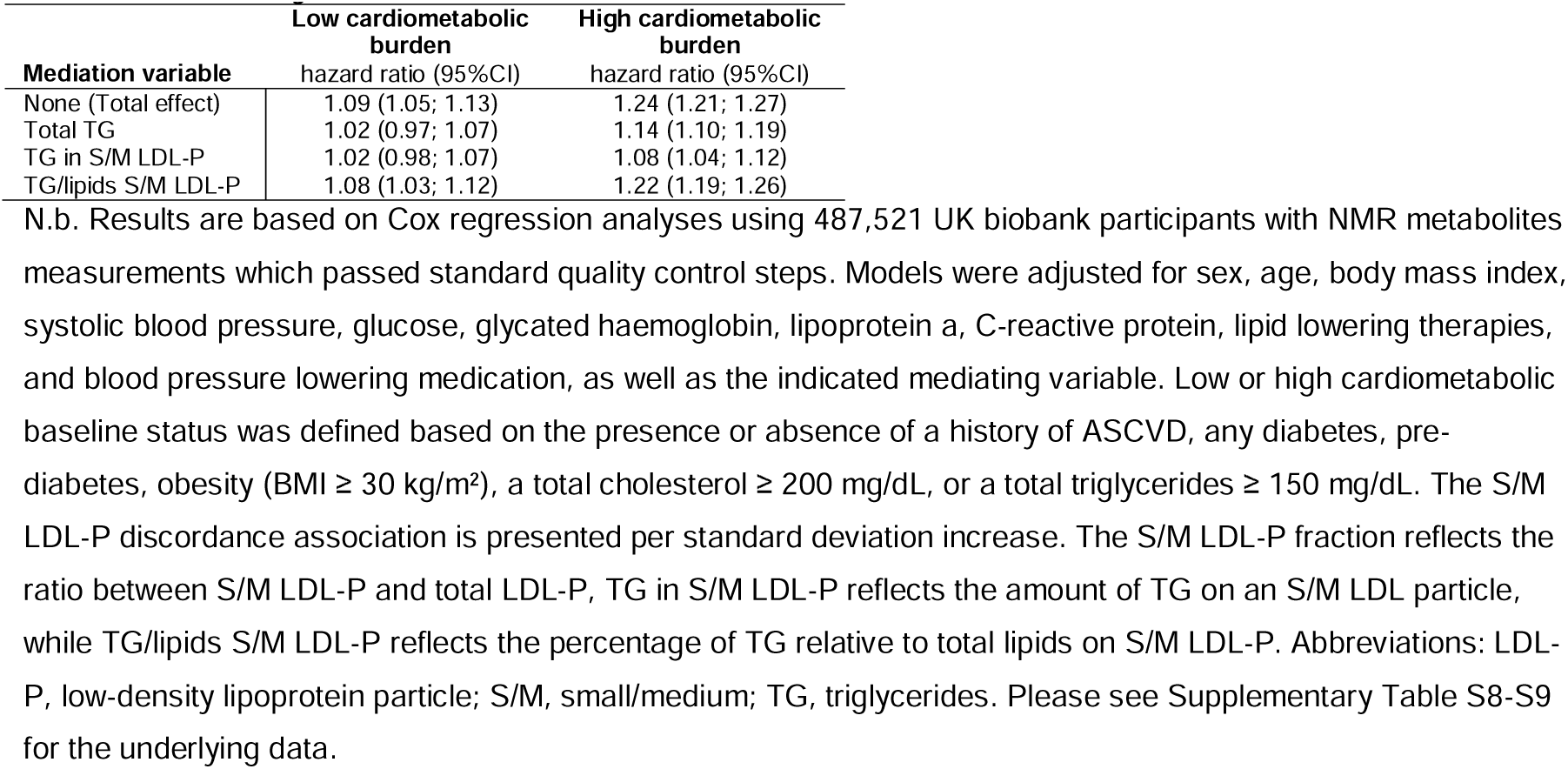
mediation analysis results estimating the direct effect of S/M LDL-P discordance on the time until major adverse cardiovascular events.

|  | Low cardiometabolic burden | High cardiometabolic burden |
| --- | --- | --- |
| Mediation variable | hazard ratio (95%CI) | hazard ratio (95%CI) |
| None (Total effect) | 1.09 (1.05; 1.13) | 1.24 (1.21; 1.27) |
| Total TG | 1.02 (0.97; 1.07) | 1.14 (1.10; 1.19) |
| TG in S/M LDL-P | 1.02 (0.98; 1.07) | 1.08 (1.04; 1.12) |
| TG/lipids S/M LDL-P | 1.08 (1.03; 1.12) | 1.22 (1.19; 1.26) |

### Sensitivity analysis

As a sensitivity analysis we estimated the correlations between the NMR and biochemistry derived measurements for LDL-C, Apo-B, and TG, which ranged between 0.80 and 0.91; Supplementary Figure S1, Supplementary Table S11. Subsequently we re-estimated S/M LDL-P discordance using biochemistry derived LDL-C “S/M LDL-P discordance (BC)”, and repeated the genetic and MACE analyses; Supplementary Table S5. The genetic associations with S/M LDL-P discordance (BC) were slightly amplified, for example *CETP* PTV carriership was associated with a mean difference change in S/M LDL-P discordance (BC) of -14.67 (-19.85; -9.50) in the high cardiometabolic burden group, this was -11.10 (-15.57; -6.63) using the original NMR derivation method; Supplementary Table S8. As before, we observed a differential MACE association based on the cardiometabolic burden group, with the S/M LDL-P discordance (BC) association limited to the higher burden group: HR per SD 1.08 (95%CI 1.06; 1.10); Supplementary Table S9.

## Discussion

In the current manuscript we created a model to estimate S/M LDL-P discordance (the difference between observed and predicted) and showed that this discordance is independent of, and thus unexplained by, LDL-C and Apo-B concentrations. Genetic variants in *CETP*, the gene encoding CETP which partially regulates TG/Cholesterol exchange and LDL particle size, were strongly associated with both S/M LDL-P itself, as well as with S/M LDL-P discordance. Genetic variants associated with increased CETP activity showed a stronger association with higher S/M LDL-P levels and discordance in people with high cardiometabolic burden, suggesting an important role of CETP in the genesis of small LDL particles among those with cardiometabolic abnormalities. By contrast *CETP* loss of function variants (a proxy for CETP-inhibiting therapies) were associated with lower S/M LDL-P discordance irrespective of metabolic burden. The association between S/M LDL-P discordance and MACE risk was most pronounced in people with high cardiometabolic burden (HR 1.24 per SD, 95%CI 1.21; 1.27). The S/M LDL-P discordance with MACE was stronger than that observed for any of the other lipid exposures considered, suggesting that conventional lipid parameters may not fully capture the atherogenic risk conferred by a greater abundance of small and medium LDL particles.

S/M LDL-P discordance was minimally associated with LDL-C and Apo-B, indicating that apparently normal levels of these standard lipid measures may mask a residual burden of atherogenic S/M LDL-P particles and the associated cardiovascular risk. Compared to S/M LDL-P concentration, S/M LDL-P discordance showed increased positive correlations with total TG, TG in S/M LDL-P, TG/lipids S/M LDL-P and S/M LDL-P fraction. This suggests that individuals with higher degrees of S/M LDL-P discordance are not only quantitatively different but also qualitatively have a distinct distribution of S/M particles which are enriched for TG content. Formal mediation analysis indicated that TG in S/M LDL-P is a strong, but possibly partial, mediator of the association between S/M LDL-P discordance with MACE. The more pronounced association between S/M LDL-P discordance and MACE among those with high cardiometabolic burden over and above S/M LDL-P concentrations per se, may therefore reflect additional qualitative changes in the particles themselves as well as other pathways correlated with discordance or potentially the longer residence time of these particles in the circulation^20,21^.

Previous studies have explored distinct types of lipid discordance, considering for example LDL-C and Apo-B or HDL-C elevation in people with normal levels of total cholesterol^22^, or between LDL-C and VLDL-C^23^. Total LDL-P discordance has previously been linked to increased insulin resistance^24^. Here we show that S/M LDL-P discordance is a more likely contributor to MACE risk in people with increased insulin resistance traits as seen among people with high cardiometabolic risk.

The following potential limitations deserve consideration. Participants were stratified according to baseline cardiometabolic burden, with the low-burden group comprising individuals free of diabetes (including pre-diabetes), established ASCVD, and obesity, while the high-burden group was correspondingly enriched for these conditions. Analyses in both groups were adjusted for key risk factors associated with higher MACE. To prevent over-adjustment, analyses were not adjusted for variables which determined cardiometabolic burden status, this would have artificially reduced the burden in the high risk group. While the current non-randomised observational analysis cannot provide proof of causality, it is important to note that the CETP inhibitor obicetrapib reduced NMR-measured small LDL-P by over 80% relative to baseline levels^25,26^. Our genetic analysis of LoF protein truncating variants in *CETP,* provides genomic evidence that potent and on-target CETP inhibition will reduce not only S/M LDL-P concentration but also S/M LDL-P discordance. Previous Mendelian randomisation studies of common genetic variants in *CETP* have consistently suggested that CETP inhibition is anticipated to reduce coronary heart disease risk^27,27,28^.

This suggests that CETP inhibition may affect MACE risk partially through a reduction of S/M LDL-P discordance. The ongoing PREVAIL trial of obicetrapib is currently evaluating its potential to meaningfully reduce MACE risk^25,26^. Irrespective of the potential causal relevance of S/M LDL-P discordance it is clear that people with elevated discordance are more likely to develop MACE and therefore require more intensive pharmacological management. The Nightingale NMR platform has consistently recapitulated established associations between NMR-derived measurements and cardiometabolic outcomes, including those relating to particle size^29,30^. It is worth flagging, however, that particle size definitions are not standardised across NMR vendors, and that the Nightingale small LDL-P measure may not capture the very smallest of LDL particles^31^. As such the reported associations may be slightly attenuated by not capturing the full spectrum of small to medium LDL particle sizes. We therefore repeated our analyses using directly measured biochemistry LDL-C, which confirmed the S/M LDL-P discordance association with genetic variants in *CETP* as well as with time till MACE onset. In this analysis the MACE association was limited to people with high baseline cardiometabolic burden, reflecting the smaller number of people with low cardiometabolic burden as well as our previous observation that S/M LDL-P is of more limited relevance in people with low cardiometabolic burden. Due to the relatively small number of people of non-European ethnicity participating in the UKB we were unable to explore potential differences due to ethnicities. Similarly, to prevent population stratification bias (where people with distinct genetic ancestry and environmental exposures are grouped together) our analyses of variants in *CETP* were limited to people of European ancestry^32^. While this does not necessarily limit the generalisability of our findings, there is a clear need to confirm effects are reasonably similar across people with different ethnicities and ancestries^15^.

In conclusion, we show that S/M LDL-P discordancy has an LDL-C and Apo-B independent association with incident MACE, which is more pronounced in people with a high baseline cardiometabolic burden. S/M LDL-P discordance is modified by loss of function genetic variation in *CETP*, suggesting a role for CETP-mediated lipid remodelling beyond LDL-C changes.

## Data Availability

The UK Biobank data used in these analyses can be requested from https://www.ukbiobank.ac.uk/, conditional on an approved research proposal.

## Funding statement

AFS is supported by BHF grant PG/22/10989, the UCL BHF Research Accelerator AA/18/6/34223, the UCL BHF Centre of Research Excellence RE/24/130013, MR/V033867/1, and the National Institute for Health and Care Research University College London Hospitals Biomedical Research Centre. This work was funded by UK Research and Innovation (UKRI) under the UK government’s Horizon Europe funding guarantee EP/Z000211/1. This publication is part of the project “Computational medicine for cardiac disease” with file number 2025.027 of the research programme “Computing Time on National Computer Facilities” which is (partly) financed by the Dutch Research Council (NWO). The authors acknowledge the use of the UCL Myriad High Performance Computing Facility (Myriad@UCL), and associated support services, in the completion of this work. MS receives salary support from CPC, a non-profit academic research organization affiliated with the University of Colorado.

## Author contributions

AFS, JK, MD designed the study. AFS performed the analyses and drafted the manuscript. NH developed the data-engineering pipelines, SQ supported the phenotyping, and MvV the genetic data extraction. AFS, NH, SQ, MvV, MdK, MS, JK, KKR, MD provided critical input on the analysis, as well as commented on the drafted manuscript.

## Conflict of Interest

AFS and MS received funding from NewAmsterdam Pharma, which is developing the CETP inhibitor obicetrapib. KKR reports unrestricted research grants (last 3 years) to Imperial College London, Amarin, Sanofi, Daiichi Sankyo, and Ultragenix; Consulting fees from Novartis, Daiichi Sankyo, Kowa, Esperion, 89bio, Novo Nordisk, MSD, Lilly, Silence Therapeutics, AZ, New Amsterdam Pharma, CLEERLY Health, SCRIBE Therapeutics, Nodthera; lecture fees from Novartis, Boehringer Ingelheim, AZ, Novo Nordisk, Amarin, Sanofi, Amgen, Daiichi Sankyo, Torrent Pharma, Algorithm, Xeedia Pharma; stock options from New Amsterdam Pharma, SCRIBE Therapeutics, Moncyte Health and PEMI 31. JK and MD are on the board of directors of NewAmsterdam Pharma. The remaining authors have no conflict of interest.

## Code availability

Analyses were conducted using Python and the packages lifelines, statsmodels, and plot-misc.

