## Supplementary Figure for "Cardiovascular events in individuals with small/medium LDL particle discordance"

**Supplementary Figure S1. The pairwise Spearman correlation of NMR and biochemistry (BC) measured lipids stratified by people with low (A) and high (B) baseline cardiometabolic burden**

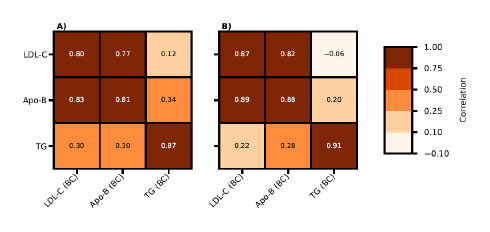

N.b. results are based on the 487,521 UK biobank participants with NMR metabolites measurements which passed standard quality control streps. Low or high cardiometabolic baseline status was defined based on the presence or absence of a history of ASCVD, any diabetes, pre-diabetes, obesity (BMI ≥ 30 kg/m²), a total cholesterol ≥ 200 mg/dL, or a total triglycerides ≥ 150 mg/dL. Abbreviations: Apo, apolipoprotein; LDL, low-density lipoprotein; TG, triglycerides. Please see Supplementary Table S11 for the underlying data.
